# The impact of damaging epilepsy and cardiac genetic variant burden in sudden death in the young

**DOI:** 10.1101/2023.03.27.23287711

**Authors:** Megan J. Puckelwartz, Lorenzo L. Pesce, Edgar J. Hernandez, Gregory Webster, Lisa M. Dellefave-Castillo, Mark W. Russell, Sarah S. Geisler, Samuel D. Kearns, Felix K Etheridge, Susan P. Etheridge, Tanner O. Monroe, Tess D. Pottinger, Prince J. Kannankeril, M. Benjamin Shoemaker, Darlene Fountain, Dan M. Roden, Heather MacLeod, Kristin M. Burns, Mark Yandell, Martin Tristani-Firouzi, Alfred L. George, Elizabeth M. McNally

**Affiliations:** Department of Pharmacology, Feinberg School of Medicine, Northwestern University, Chicago, IL; Center for Genetic Medicine, Feinberg School of Medicine, Northwestern University, Chicago, IL; Biomedical Informatics, University of Utah, Salt Lake City, Utah; Division of Cardiology, Department of Pediatrics, Ann & Robert H. Lurie Children’s Hospital of Chicago, Chicago, IL; Department of Pediatrics, University of Michigan, Salt Lake City, Utah; Division of Pediatric Cardiology, University of Utah, Salt Lake City, Utah; Department of Pediatrics, Vanderbilt University Medical Center, Nashville, TN; Department of Medicine, Division of Cardiovascular Medicine, Vanderbilt University Medical Center, Nashville, TN; Departments of Medicine and Pharmacology, Vanderbilt University Medical Center, Nashville, TN; Data Coordinating Center, SDY Case Registry, Bethesda, Maryland; Division of Cardiovascular Sciences, National Heart, Lung, and Blood Institute, National Institutes of Health, Bethesda, Maryland; Department of Human Genetics, University of Utah, Salt Lake City, Utah

**Keywords:** sudden death in the young, genome sequencing, epilepsy, arrhythmia, cardiomyopathy, gene burden

## Abstract

**Background:** Sudden unexpected death in children is a tragic event. Understanding the genetics of sudden death in the young (SDY) enables family counseling and cascade screening. The objective of this study was to characterize genetic variation in an SDY cohort using whole genome sequencing.

**Methods:** The SDY Case Registry is a National Institutes of Health/Centers for Disease Control surveillance effort to discern the prevalence, causes, and risk factors for SDY. The SDY Case Registry prospectively collected clinical data and DNA biospecimens from SDY cases <20 years of age. SDY cases were collected from medical examiner and coroner offices spanning 13 US jurisdictions from 2015-2019. The cohort included 211 children (mean age 1 year; range 0-20 years), determined to have died suddenly and unexpectedly and in whom DNA biospecimens and next-of-kin consent were ascertained. A control cohort consisted of 211 randomly sampled, sex-and ancestry-matched individuals from the 1000 Genomes Project. Genetic variation was evaluated in epilepsy, cardiomyopathy and arrhythmia genes in the SDY and control cohorts. American College of Medical Genetics/Genomics guidelines were used to classify variants as pathogenic or likely pathogenic. Additionally, genetic variation predicted to be damaging was identified using a Bayesian-based artificial intelligence (AI) tool.

**Results:** The SDY cohort was 42% European, 30% African, 17% Hispanic, and 11% with mixed ancestries, and 39% female. Six percent of the cohort was found to harbor a pathogenic or likely pathogenic genetic variant in an epilepsy, cardiomyopathy or arrhythmia gene. The genomes of SDY cases, but not controls, were enriched for rare, damaging variants in epilepsy, cardiomyopathy and arrhythmia-related genes. A greater number of rare epilepsy genetic variants correlated with younger age at death.

**Conclusions:** While damaging cardiomyopathy and arrhythmia genes are recognized contributors to SDY, we also observed an enrichment in epilepsy-related genes in the SDY cohort, and a correlation between rare epilepsy variation and younger age at death. These findings emphasize the importance of considering epilepsy genes when evaluating SDY.

## BACKGROUND

Sudden death in children has immediate and sustained medical and emotional consequences for affected families. Elucidating the etiology of sudden death can inform risk for surviving family members. The Sudden Death in the Young (SDY) Case Registry was created as a joint initiative between the National Institutes of Health (NIH) and the Centers for Disease Control and Prevention (CDC) to enable a population-based surveillance of SDY and facilitate research through the collection of clinical data and biospecimens (1). The SDY Case Registry engaged public health agencies in thirteen diverse US sites to collect information on children, ages 0-20 years, who died suddenly and unexpectedly. Data from the SDY Registry indicates that sudden death mortality rates are greater for infants less than 1 year (120/100000 live births) than for children 1-17 years (1.9/100000 children) (2). SDY can be attributed to primary cardiac causes including arrhythmias, cardiomyopathies, congenital heart defects, and vascular disease, as well as noncardiac causes such as epilepsy. While extensive work has established social and environmental factors that contribute to sudden infant death syndrome (SIDS), the genetic contributors to sudden unexpected infant death (SUID), sudden cardiac death, and sudden unexpected death in epilepsy (SUDEP) remain incompletely described.

There is increasing evidence that SDY has a genetic component and postmortem genetic screening can aid in identifying the cause of death. Testing of genes implicated in cardiac rhythm and function identifies a pathogenic or likely pathogenic (P/LP) variant in 10-25% of individuals with sudden unexplained death <40 years old (3, 4). Approximately 4% of SIDS may be due to clinically actionable genetic cardiac causes (5). In a cohort of sudden unexpected death in pediatrics (SUPD), contributory genetic variants were identified in 11% of decedents, and the authors also found an excess burden of rare, damaging SUDEP gene variants compared to a control group (6). We previously used genome sequencing (GS) to analyze an SDY cohort (n=103) covering a larger age range (1-44 years) and found ∼13% carried a pathogenic/likely pathogenic (P/LP) variant in an arrhythmia or cardiomyopathy gene. Younger decedents carried an excess of suspicious variants of uncertain significance (VUS) and P/LP variants compared to a control population. Furthermore, for decedents >2 years of age at death, a younger age was associated with harboring more rare cardiac variants (7). Here, we interrogated the genomes of 211 decedents (≤19 years old) ascertained through the NIH/CDC SDY Registry with a median age at death of 1 year, with an analysis of genes associated with epilepsy, cardiomyopathies and arrhythmias.

## METHODS

### SDY Case Registry

The SDY Case Registry was previously described (1). Briefly, the SDY Case Registry aggregated information from 13 participating states and jurisdictions in collaboration with public health agencies, including medical examiner offices. Clinical data were collected via the US National Child Death Review Case Reporting System. The Institutional Review Board (IRB) at the Office of Research Integrity at the Michigan Public Health Institute (MPHI) provided ethical approval for the study (initial approval 2015 with subsequent annual renewals). The MPHI was the central IRB, and for those jurisdictions who did not rely on the Central IRB, local IRBs provided approval for the study. Parents/next of kin provided written informed consent for research use of the decedent’s DNA and linked information with consent to broad, future, unspecified research on their child’s DNA and linked information to create a research repository. The MPHI also served as the Data Coordinating Center (DCC) for the SDY Case Registry. Biospecimens for DNA extraction were obtained at autopsy. All data was deidentified for the analysis performed in this work.

#### Genome Sequence (GS) Analysis

*Sequencing methods are detailed in the Supplement.* Variants were called with either the Genome Analysis Tool Kit (GATK v3.3.0) using the MegaSeq pipeline or Sentieon joint variant calling software (8, 9, 10). Variant call files were annotated using SnpEff (11). Variants were annotated using ClinVar (accessed March 2022) and global and ancestry-specific allele frequencies using the Genome Aggregation Database (gnomAD) (12). M-CAP was used to annotate variant pathogenicity (13).

#### Genetic Evaluation of Ancestry

Ancestry was determined using principal component analysis (PCA) conducted using singular-value decomposition of ∼5 million biallelic variants across the genome. PCAs were generated using the first 2 components for global ancestry estimates. Asian ancestry was determined using the 3^rd^ PC. Analyses were performed using PLINK v1.9 and R v4.1.

#### Gene Panel Analysis

Four previously-established gene panels were analyzed (two epilepsy panels and two arrhythmia and cardiac function panels, eTables 2 and 3 in the Supplement): 1) The Early Infantile Epileptic Encephalopathy (EIEE)- Online Mendelian Inheritance in Man (OMIM) panel (82 genes) derived from the OMIM phenotypic series for early infantile epileptic encephalopathy, a curated epilepsy list; 2) The Epilepsy gene panel (191 genes, overlaps with the EIEE panel), and is a curated list derived from the Invitae Epilepsy Panel (Invitae, San Francisco, CA); 3) CMAR1, CardioMyopathy and Arrhythmia (gene panel including 118 arrythmia and cardiac genes previously described (7); and 4) CMAR2, a Pan-Cardiomyopathy Arrhythmia and Cardiomyopathy Comprehensive gene panel including 143 genes with overlap with the CMAR1 panel (14). Control genes were selected from uniformly distributed housekeeping genes (15, 16). To create burden-matched control gene lists, a burden ratio measurement was calculated for each gene in RefSeq by counting the number of rare variants (MAF ≤ 0.005) found in the gnomAD database over the largest coding transcript of every gene divided by transcript length (12). The mean burden ratio and standard deviation from each list were matched by randomly sampling from the whole genome (excluding genes in the test list) (15, 17).

#### Artificial-Intelligence (AI)-Based Variant Prioritization

AI-based prioritization and scoring of candidate disease genes and diagnostic conditions were performed using GEM (18) in the commercially available Fabric Enterprise platform (Fabric Genomics, Oakland, CA). Briefly, GEM aggregates data from multiple sources and clinical data sets to identify pathogenic variants relevant to phenotype terms input by the user. The GEM pipeline requires a variant call file, affection status, and human phenotype ontology (HPO) terms for each individual. A log_10_ Bayes-factor score (GEM score) was generated that calculated the extent of support for a given model using multiple lines of evidence from open-source tools and databases. A GEM score ≥0.69 was used to define a likely deleterious genetic variant, based on the observation this threshold recovered 95% of true positive cases in a cohort of critically ill newborns undergoing rapid genome sequencing (18).

#### Control Cohort

The control cohort of 211 genome samples derived from the 1000 Genomes Project (https://www.internationalgenome.org/) that were variant called using the Sentieon pipeline described above against the GRCh37/hg19 human reference genome(19). This control cohort matched the ancestry and sex composition as the SDY cohort.

#### Enrichment Analysis

Since detailed clinical information was lacking from the SDY cohort, we used HPO terms expected to be associated with SDY: Sudden Cardiac Death (HP:0001645), Sudden Death (HP:0001699), Cardiac arrest (HP:0001695), Cardiomyopathy (HP:0001638), Abnormal QT interval (HP:0031547), and Seizure (HP:0001250). We also performed the analysis using a control root phenotype, Phenotypic Abnormality (HP:0000118) which was used as a normalization factor to control for ontology biases and was expected to behave similarly across different cohorts.

GEM analysis was performed for each HPO term with each iteration identifying potentially causal variants for the three phenotypes described. To assess enrichment between cases and the controls a resampling analysis was performed as previously described (17). Briefly, for each investigated gene list (CMAR1, CMAR2, EIEE-OMIM, and Epilepsy Panels) of size N, 100,000 random samples of equal size were drawn from the 18,876 RefSeq genes and intersected with the corresponding GEM list of damaged genes producing the number of damaged genes in the resampled list. Fisher exact test was used to determine if there were significant differences in the number of genes between cases and controls using the permutation test to normalize the data across runs and produce an empirical p-value (17). Multiple comparison correction was performed using the False Discovery Rate (FDR) approach (20). Each null distribution was scaled and centered producing Z-scores to directly compare the p values of different gene sets.

#### Age at death burden analysis

Age at death dependence was correlated with the cumulative number of nonsynonymous variants in the Epilepsy or CMAR gene panels. Variants were binned by global gnomAD frequency [<0.001, 0.001-0.01, 0.01-0.1, 0.1-0.25, 0.25-0.5]. Multivariate linear models adjusting for ancestry (PCs 1-6) using custom code based on the *lm* function in R 4.1 were regressed to test each bin. P-values were adjusted for multiple comparisons. An extreme value sensitivity test was performed for decedents at the ends of the rare variant distribution (gnomAD <0.001); p values were adjusted for ancestry PCs 1-6.

#### Variants of Uncertain Significance (VUS) Analysis

The number of nonsynonymous, rare variants (<0.001) found in gnomAD (V2) for each gene in the Epilepsy and CMAR1 gene panels was compiled. To determine expected number of variants in each gene for the gene set used (Epilepsy/CMAR), we created a multinomial model using the observed number of missense variants in the same gene set in gnomAD. We estimated the probability of observing a variant in each gene in the gene set as the observed number of variants in gnomAD by dividing the total number of variants in the gene set in gnomAD.

## RESULTS

### The Sudden Death in the Young (SDY) Cohort

In total, 230 decedents from the SDY registry had full genome sequencing (**Figure 1**). Of the 230, 19 were excluded; 4 due to failed inclusion criteria and 15 due to failed sequencing where no additional sample was available. Of the remaining 211, 152 (72%) had partial phenotype data available from the SDY Case Registry (**Table 1**). The median age at death was 1 year [0.33, 0.96] (median, IQR), indicating an enriched sample of cases who died in the first year of life. Genetic ancestry analysis was used to classify SDY decedents as: 42% European, 30% African, 17% Hispanic, and 11% with mixed ancestries. The cohort was 61% male and 39% female (**Table 1**). The SDY Case Registry provided cause of death information by standardized case report forms. The cause of death was included for 152 decedents, including 104 whose death remained unexplained and 48 with a known cause of death reported. Detailed cause of death is noted in **eTable 1** in the Supplement with sudden unexplained death as the most common cited cause (43%).

**Figure 1.**
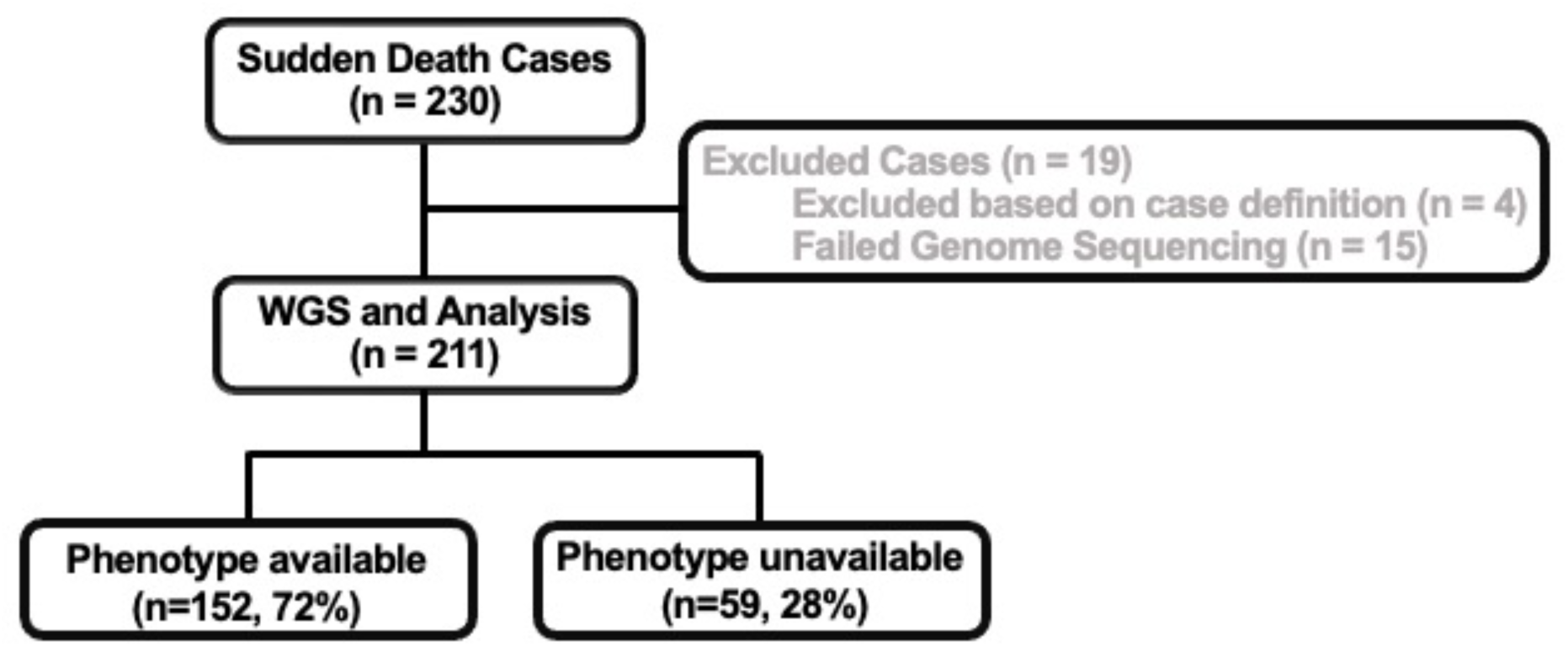
CONSORT diagram of decedents selected for genome sequencing. GS = genome sequencing.

**Table 1.** Demographics of study cohort (n=211)

| **Phenotype Information** | **Yes** | **No** |
| --- | --- | --- |
| **n (%)** | 152 (72) | 59 (28) |
| **Age at death (median [IQR])** | 1 [0.33,0.96] | NA |
| **Genetic Ancestry (%)** |  |  |
| **African** | 50 (32.9) | 13 (22) |
| **European** | 68 (44.7) | 21(35.6) |
| **Hispanic** | 21 (13.8) | 15(25.4) |
| **Mixed** | 13 (8.6) | 10(16.9) |
| **Genetic Sex (% male)** | 92 (60.5) | 36 (61.0) |

### Pathogenic/likely pathogenic variants in cardiac and epilepsy genes

To identify variants that may have contributed to SDY, we employed gene panels to filter for rare, high impact variants. We hypothesized that the most likely candidates for SDY would be epilepsy, and cardiomyopathy and arrhythmia gene variants. We identified nonsynonymous genetic variants in 161 out of 191 genes for the Epilepsy panel and in 99 out of 118 of the CMAR1 genes (**eTable 2** and 3 in the Supplement). Variants were annotated using ACMG classification and ClinVar interpretations to designate variants as pathogenic (P), likely pathogenic (LP), or variant of uncertain significance (VUS).

Within the two gene sets we identified P/LP variants in ∼6% of decedents (11 of 211). In these 11 individuals, the same *TTR* variant appeared in 4 decedents (**eTable 4** in the Supplement). In the epilepsy panel, we identified 4 variants in 3 decedents, but these genes generally associate with autosomal recessive inheritance. Six individuals had at least one P or LP variant in genes from the CMAR1 panel. One variant was identified in the *FKRP* gene which associates with recessive disease. One individual had a variant in the *KCNH2* gene, which is found in both the Epilepsy and CMAR1 panels (**eTable 4** in the Supplement).

To identify potentially pathogenic variants in a broader set of genes without constraining to the Epilepsy and CMAR panels, we deployed GEM, an AI tool designed for whole genome-based diagnosis of Mendelian conditions (18), using HPO terms related to seizures and sudden cardiac death. Thirty-eight decedents (18%) harbored ClinVar-designated P/LP variants in genes linked to a Mendelian disorder (13 related to the HPO term seizure and 25 related to sudden cardiac death; **eTable 5** in the Supplement). Four of these decedents harbored compound heterozygous damaging variants in genes associated with recessive disorders, including *CFTR* (Cystic Fibrosis), *BCDHA* (Maple Syrup Urine Disease), *MPDZ* (Congenital Hydrocephalus-2), and *GDAP1* (Charcot-Marie-Tooth Disease, Type 4A).

### Damaging Genetic Variation in Cardiac and Epilepsy Genes was Enriched in the SDY cohort

We compared the burden of damaging cardiac and epilepsy genes in SDY cases versus controls, using two CMAR and two Epilepsy gene panels (see Methods). A GEM score ≥0.69 was used to define a likely deleterious genetic variant (see Methods). We also used a known housekeeping gene list and a random sample of genes matched for similar variant burden (16). **Figure 2** provides the distributions for the random gene samples (gray bars) and the number of genes identified for enrichment in the gene lists (dark arrows) and the burden-matched controls (light arrows). This analysis revealed an increased burden of genes with damaging variants for both Epilepsy and CMAR1 panels when compared with the control gene set (**Figure 2A** and **B**, respectively). A similar pattern was seen when we analyzed the EIEE-OMIM and CMAR2 gene panels (**eFigure 1** in the Supplement). The Fisher exact-test p-values FDR adjusted are Epilepsy p<0.027, EIEE-OMIM p<0.019, CMAR1 p<0.001, CMAR2 p<0.001 (**eTable 6** in the Supplement).

**Figure 2.**
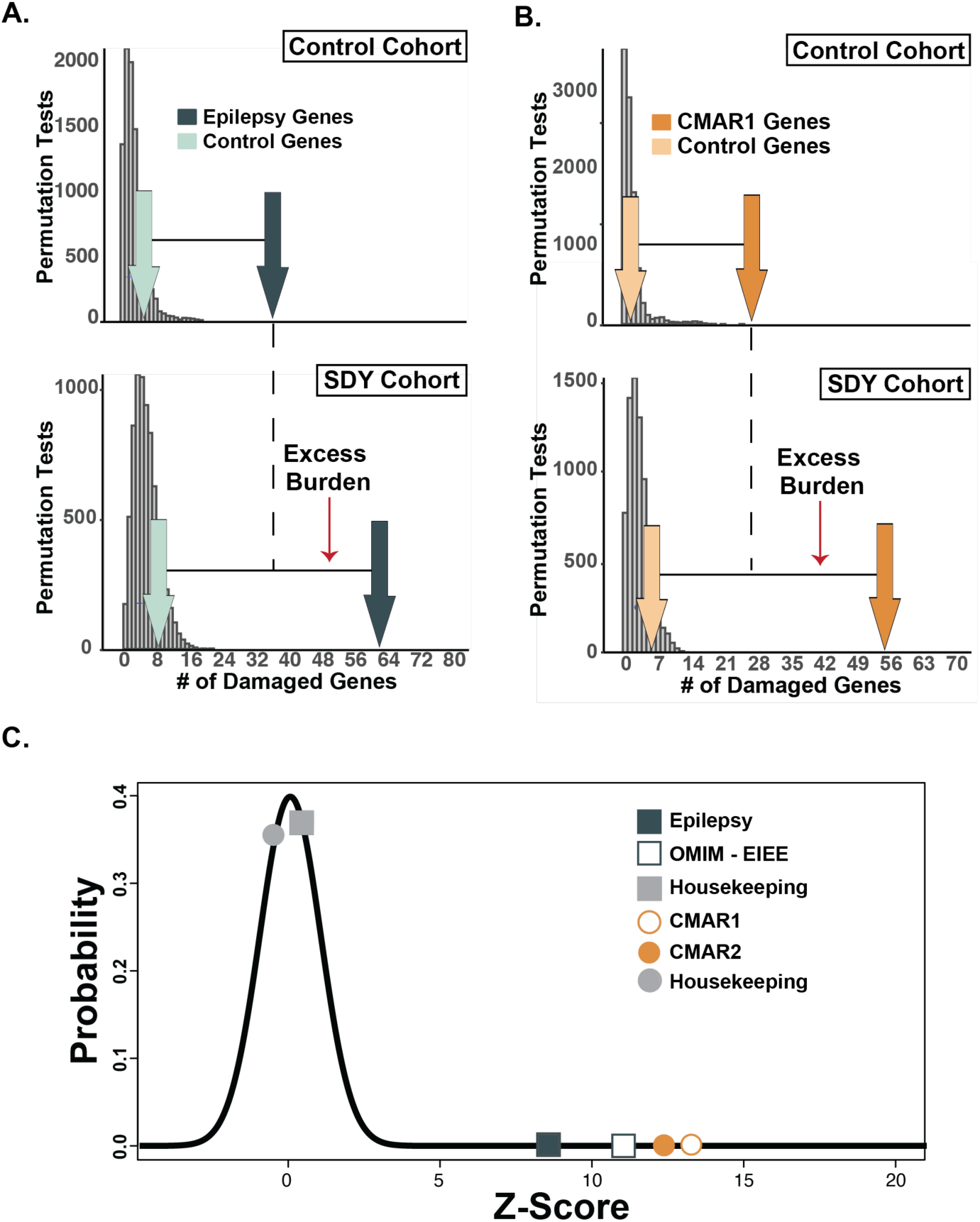
**Relative enrichment of damaging variants in epilepsy and Cardiomyopathy/Arrhythmia (CMAR) genes.** The SDY cohort has enriched damaging (A) epilepsy and (B) cardiac gene burden compared to a sex-and ancestry-matched control cohort. Histograms (gray bars) represent distributions of damaged genes (GEM Score >0.69) sampled randomly from RefSeq genes using a root phenotype from the SDY cohort (n=211) (bottom panels) and 1000 Genomes Project Cohort (control) matched for sex and ancestry (n=211) (top panels). Damaged genes identified in the Epilepsy (n=191 genes, green) **(A)** and CMAR1 (n=118 genes, orange) **(B)** gene lists were significantly different between the SDY and control cohorts (dark arrows, epilepsy, p=0.027; cardiac p<0.001). Light arrows represent the number of damaged genes identified in a control housekeeping gene set. **(C)** The results in Figure 1A and B and eFigure 1 were Z-score transformed to make the gene list findings comparable. The plot reveals a considerable enrichment in both cardiac and epilepsy gene lists (+8.0- +13.0 SDs). Black line represents the expected normal distribution.

To compare the findings among the distributions shown in **Figure 2A** and **B**, each distribution was normalized by Z-score transformation (**Figure 2C**). The standardized Z-score values of the enrichment observed in the gene panels (**Figure 2C**) exceeded the expected normal distribution (shown in black), indicating a strong enrichment for damaging variation in Epilepsy and CMAR genes in the SDY cohort (+8.0-+13.0 standard deviations).

### Rare Variants in Epilepsy Genes Correlated with Age at Death

The enrichment of predicted damaging variation in Epilepsy and CMAR genes and the few decedents with P/LP variants, led us to determine if there is an association between rare Epilepsy and CMAR variants and age at death. We aggregated variants across five gnomAD allele frequency bins (<0.001, 0.001-0.01, 0.01-0.1, 0.1-0.25, 0.25-0.5) for the Epilepsy and CMAR1 gene panels. We found that having more rare, nonsynonymous variants in Epilepsy genes was associated with younger age at death. Nonsynonymous Epilepsy variants with an allele frequency <0.001 were significantly associated with age at death, (p=0.0053, adjusted for ancestry) (**Figure 3A** and **eTable 7** in the Supplement). Results remained significant (p=0.026) following correction for multiple hypothesis testing for all 5 frequency bins. To confirm that these data were not driven by a few outliers, we performed a sensitivity analysis that supported the results of the main analysis (**eTable 7** in the Supplement). A similar analysis of rare, nonsynonymous variants in the CMAR1 gene panel did not show an association for age at death in any frequency bin (**Figure 3B**).

**Figure 3.**
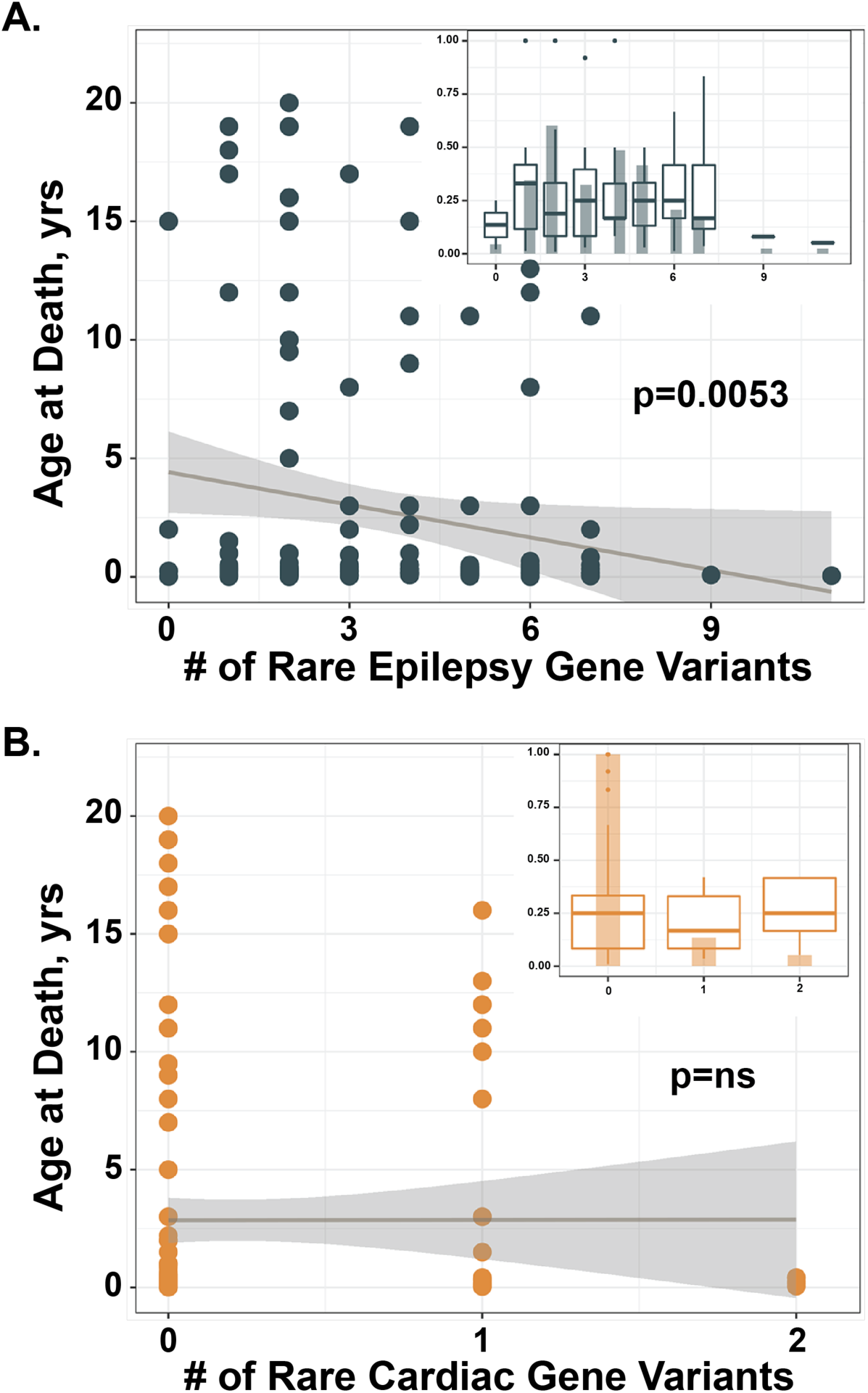
**Number of rare, nonsynonymous, epilepsy gene variants correlated with younger age at death**. The age at death was plotted against against the number of rare variants (gnomAD allele frequency <0.001) in either the **(A)** Epilepsy or **(B)** CMAR1 gene list. Number of variants in the epilepsy gene list significantly correlated with age at death, (p=0.0053, adjusted for the first 6 principal components of ancestry). A similar analysis of rare, predicted damaging variation in the CMAR1 genes did not show association with age at death (p=0.85, p value adjusted for ancestry). Inset is a histogram showing number of decedents <1 year old by variant burden, box plots (black) represent decedent age distribution for each variant burden. Dark gray line = regression line of unadjusted model; dark gray shading represents 95% confidence intervals.

## Rare VUS in Epilepsy and Cardiac Genes were enriched in the SDY cohort

We next considered if variants of uncertain significance (VUS) might associate with SDY. VUS are rare variants about which there is insufficient information as to whether they are pathogenic or benign, and the major determinant of VUS status is rare population frequency. **Figure 4** plots the number of rare, nonsynonymous variants (<0.001 gnomAD allele frequency) not reported as pathogenic or benign in ClinVar. Variants were scored using the *in silico* predictor M-CAP and scored as damaging using an M-CAP score >0.025. We plotted total variants, damaging variants, expected number of variants based on gnomAD distribution and age at death (3^rd^ quartile) of each decedent, and we found that genes in both Epilepsy and CMAR1 panels had greater than expected numbers of variants. Some genes had very few damaging variants, while others had more than expected. Those genes with greater than expected variation may potentially drive or modify risk for SDY.

**Figure 4.**
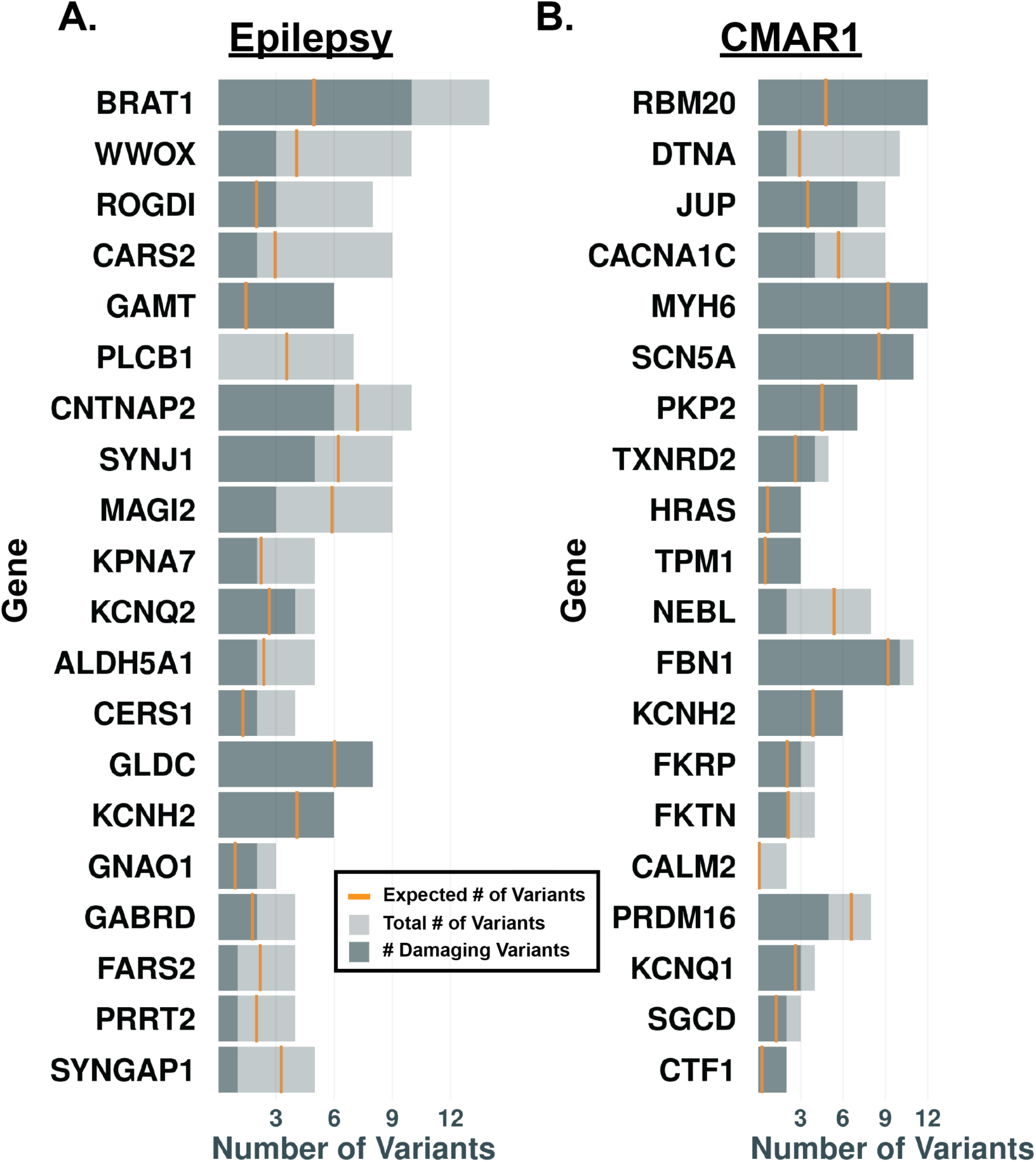
**Rare VUS in Epilepsy and CMAR Genes.** The number of rare (<0.001 gnomAD allele frequency) variants not classified as pathogenic or benign in ClinVar are reported for genes from either the **(A)** Epilepsy or **(B)** CMAR1 gene list (light gray bars). Variants were scored using the *in silico* tool M-CAP. The number of damaging VUS for each gene (>0.025 M-CAP score) is shown by the dark gray bars. The gold line indicates the expected number of variants based on gnomAD allele frequency data. Gene lists are ranked by the difference between the observed and expected number of variants, when ratios are approximately the same, ranking is determined by number of damaging gene variants.

## DISCUSSION

This SDY cohort is unique for its very young age (mean age 1 year) and its diverse ancestry. In the SDY cohort, we identified P/LP variants in Epilepsy and CMAR genes in ∼6% of the cohort. An unrestrained AI-driven analysis identified 42 P/LP variants in 38 decedents suggesting that a purely gene panel-based approach may be diagnostically limited. Previous studies identified that the prevalence of P/LP gene variants is less in younger cohorts (3, 4, 5, 21), consistent with findings in our young cohort. In adults, the prevalence is likely 15-20%; whereas, in infants, our data agrees with others that the prevalence is < 10% when cardiomyopathy and arrhythmia genes are considered with a panel approach. Analysis of 278 SIDS cases for ultrarare variants in noncardiac, SIDS-susceptibility genes did not identify a monogenic basis for SIDS, even when using pathway burden analyses (22).

A recent study of SDY examined decedents >1 to 44 years of age, and specifically excluded the group under 1 year of age (7). A similar burden analysis found more CMAR1 gene variants and an association between gene variant burden and younger age at death (7). There is evidence that epilepsy-related mechanisms associate with SDY pathogenesis (23, 24). This younger SDY cohort, with a median age of 1 year at time of death, displayed enrichment of both epilepsy and cardiac damaging variants, but only the epilepsy gene variant burden correlated with younger age at death. These data are in line with another study showing an increased burden in epilepsy, cardiac and metabolic genes with an excess of rare variants in all three gene classes in SUDP cases ≤1 year old (6).

## CONCLUSIONS

Together, these findings support epilepsy etiologies for SDY, particularly in infants with a broader genetic contribution including cardiac and epilepsy variants in those >1 year of age. Even more relevant, the genetic associations for SDY appear to derive less from single gene P/LP variants and instead correlate with an aggregation of potentially damaging variants within an individual genome that predispose to SDY.

## Data Availability

Both phenotype (where available) and sequence data will be available via dbGAP after publication or by reasonable request to the corresponding author. Relevant data and scripts will be made available upon request to the corresponding author.

## LIST OF ABBREVIATIONS

*SDY*: Sudden death in the young
NIH: National Instistutes of Health
CDC: Center for Disease Control
SIDS: Sudden infant death syndrome
SUID: Sudden unexprected infant death
SUDEP: Sudden unexplained death in epilepsy
P/LP: Pathogenic or likely pathogenic
SUDP: Sudden unexpected death in pediatrics
GS: Genome sequencing
VUS: Variant of unknown significance
gnomAD: Genome aggregation database
PCA: Principal component analysis
EIEE: Early infantile epileptic encephalopathy
OMIM: Online Mendelian inheritance in man
CMAR1: CardioMyopathy and Arrhythmia panel 1
CMAR2: CardioMyopathy and Arrhythmia panel 2
AI: Artificial intelligence
HPO: Human phenotype ontology
FDR: False discovery rate

## DECLARATIONS

### ETHICS APPROVAL AND CONSENT TO PARTICIPATE

We attest that data included in this manuscript was conducted in a manner consistent with principles of research ethics per the standards of the Belmont Report. This research was performed with the voluntary, informed consent of the next of kin of the decedent, free from coercion. Registry activities involving biospecimen collection and consent of surviving family members for research were approved by the institutional review boards at the Data Coordinating Center and participating states/jurisdictions.

### CONSENT FOR PUBLICATION NA

#### AVAILABILITY OF DATA AND MATERIALS

Both phenotype (where available) and sequence data will be available via dbGAP after publication or by reasonable request to corresponding author. Relevant data and scripts will be made available upon request to the corresponding author.

## AUTHOR CONTRIBUTIONS

MJP, GW, LMDC, KMB, MTF, ALG, and EMM conceptualized the project. MJP, LMM, EJH, GW,SDK, MY developed methods including scripts for data analysis. MJP, LLP, GW, LMDC, SDK, FKF, HM, KMB participated in data curation. MJP, LLP, EJH, performed formal analysis of the data. MJP, LLP, EJH, GW, LMDC, FKF, HM, KMB conducted experiments through data collection and in silico analysis. TOM, TDP validated data and experiments. MJP was responsible for the first draft of the manuscript. LLP, EJH, GW, LMDC, SPE, PJK, MBS, DF, DMR, KMB, HM, MY, MTF, ALG and EMM carefully reviewed data and experiments and edited the manuscript. PJK, HM, KMB coordinated collection of decedent samples and information. MJP, PJK MTF, ALG and EMM secured funding.

## ACKNOWLEDGEMENTS

The views expressed in this manuscript are those of the authors and do not reflect official positions of the National Heart, Lung, and Blood Institute, the National Institutes of Health, or the United States Department of Health and Human Services.

This work used Stampede2 at The Texas Advanced Computing Center (TACC) through allocation XRAC - BIO220012 (PI MJP) from the Extreme Science and Engineering Discovery Environment (XSEDE), which was supported by National Science Foundation grant number #1548562 (25). We thank Victor EijKhout for his assistance with porting and optimization, which was made possible through the XSEDE Extended Collaborative Support Service (ECSS) program (26). We also thank Justin Wozniak for his support in installing and optimizing our swift-t workflow (27).

*The information that is the basis of this presentation and/or publication was provided by the NCRPCD/DCC, which is funded in part by the US Centers for Disease Control and Prevention Division of Reproductive Health (CDC) and the Health Resources and Services Administration. The data is part of the Sudden Death in the Young (SDY) Case Registry, which is funded by the National Heart, Lung, and Blood Institute and the National Institute of Neurologic Disorders and Stroke of the National Institutes of Health (NIH) and the Centers for Disease Control and Prevention. The contents are solely the responsibility of the authors and do not necessarily represent the official views of NCRPCD, CDC, NIH or the participating states*.

## SUPPLEMENT

### Supplemental Methods

WGS was performed on DNA obtained from the SDY Case Registry using an Illumina XTen sequencer (Garvan Institute of Medical Research NSW, Australia) with a yield of >100 GB sample, correlating to >30-fold coverage across the genome. The Burrows- Wheeler Aligner was used to align reads to human reference sequence GRCh37/hg19 (28).

**Supplemental Figure 1.**
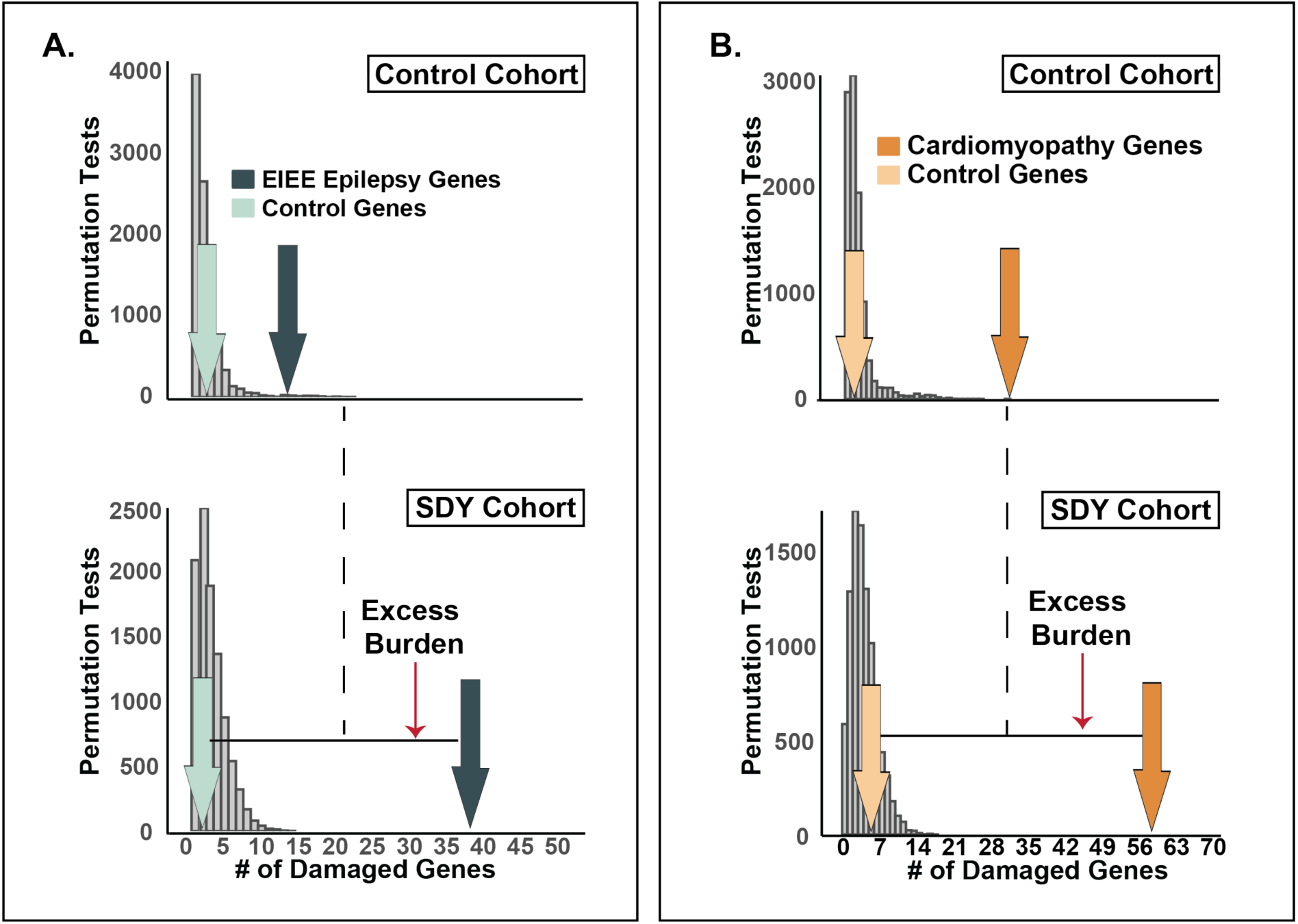
The SDY cohort had enriched damaging (A) epilepsy and (B) CMAR2 gene burden compared to a sex- and ancestry-matched control cohort. Histograms (gray bars) represent distributions of damaged genes (GEM Score >0.69) sampled randomly from RefSeq genes using a root phenotype from the SDY cohort (n=211) (bottom panels) and 1000 Genomes Project Cohort (control) matched for sex and ancestry (n=211) (top panels). Damaged genes identified in the Epilepsy EIEE (n=82 genes, green) (A) and CMAR2 (n=143 genes, orange) (B) gene lists were significantly different between the SDY and control cohorts (dark arrows, epilepsy, p=0.019; cardiac p<0.001). Light arrows represent the number of damaged genes identified in a control gene set.

**Supplemental Table 1.** Detailed Cause of Death

| **Cause of Death** | **Number of Decedents (%)** |
| --- | --- |
| **Unexplained (n=104, 49%)** |  |
| Sudden unexplained death | 90 (43) |
| Possible SUDEP | 6 (3) |
| Cardiac findings of uncertain significance | 5 (2) |
| Concerning family history | 3 (1) |
| **Explained (n=48, 23%)** |  |
| Infant suffocation | 15 (7) |
| Systemic viral or bacterial infection | 13 (6) |
| Asthma | 3 (1) |
| Cardiomyopathy | 3 (1) |
| Drowning | 3 (1) |
| Multisystem failure in complex medical patient | 3 (1) |
| Major vascular malformation | 2 (1) |
| Myocarditis | 2 (1) |
| Congenital heart disease | 2 (1) |
| Diabetic ketoacidosis | 1 (<1) |
| Pulmonary embolism | 1 (<1) |
| No phenotype information available | 59 (28) |
| **TOTAL** | **211** |

SUDEP = Sudden Unexplained Death in Epilepsy. Cardiac findings of uncertain significance included findings of cardiomyopathy without sufficient criteria for diagnosis (2 decedents), possible coronary artery disease (2), and aortic valve abnormality with left ventricular hypertrophy (1). Concerning family history included long QT syndrome, sudden unexplained death in a sibling, and family history of heart attack and stroke < 50 years of age, each in 1 decedent. Congenital heart disease included hypoplastic left heart syndrome (1) and anomalous left coronary artery (1).

**Supplemental Table 2.**
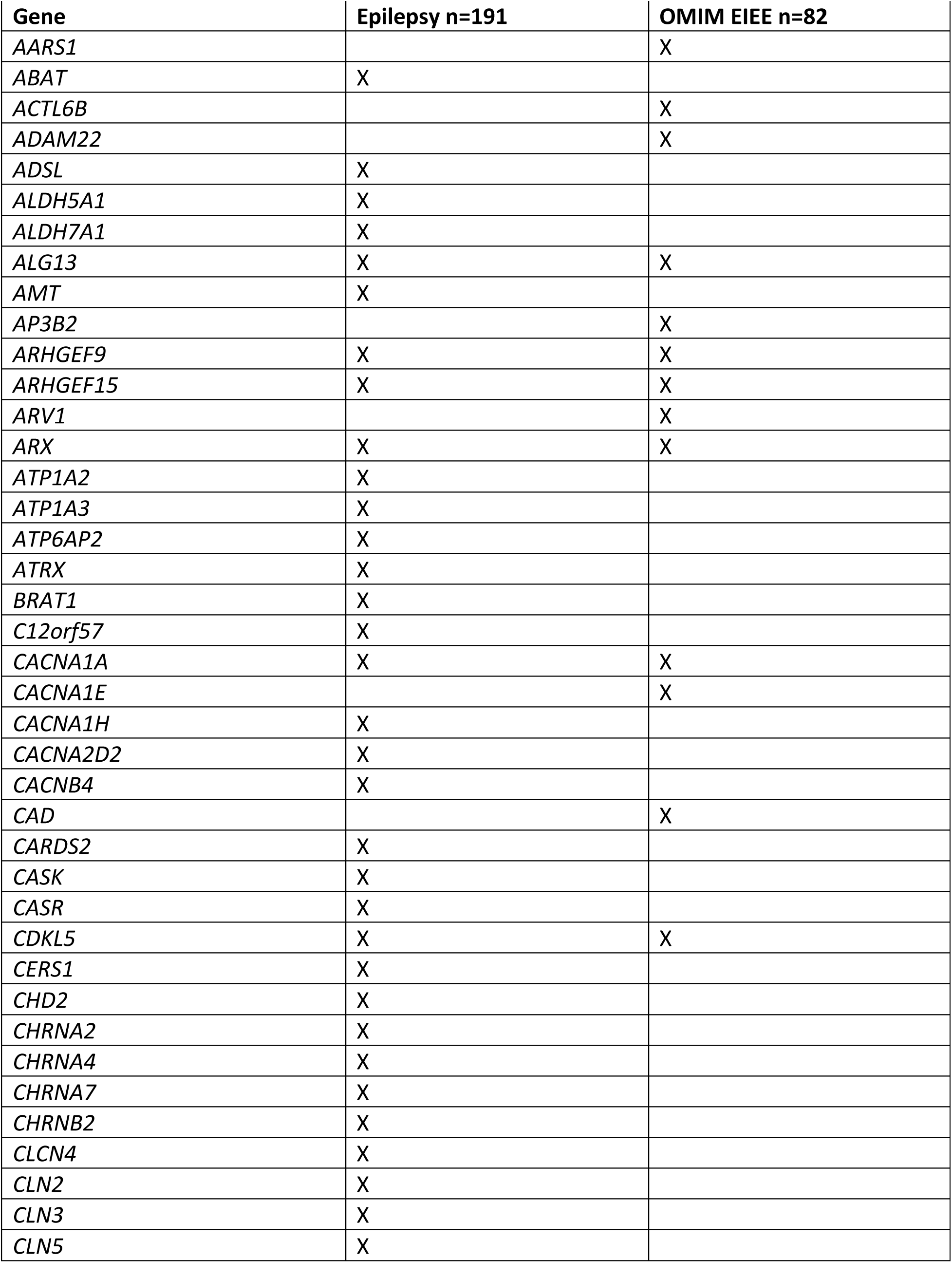
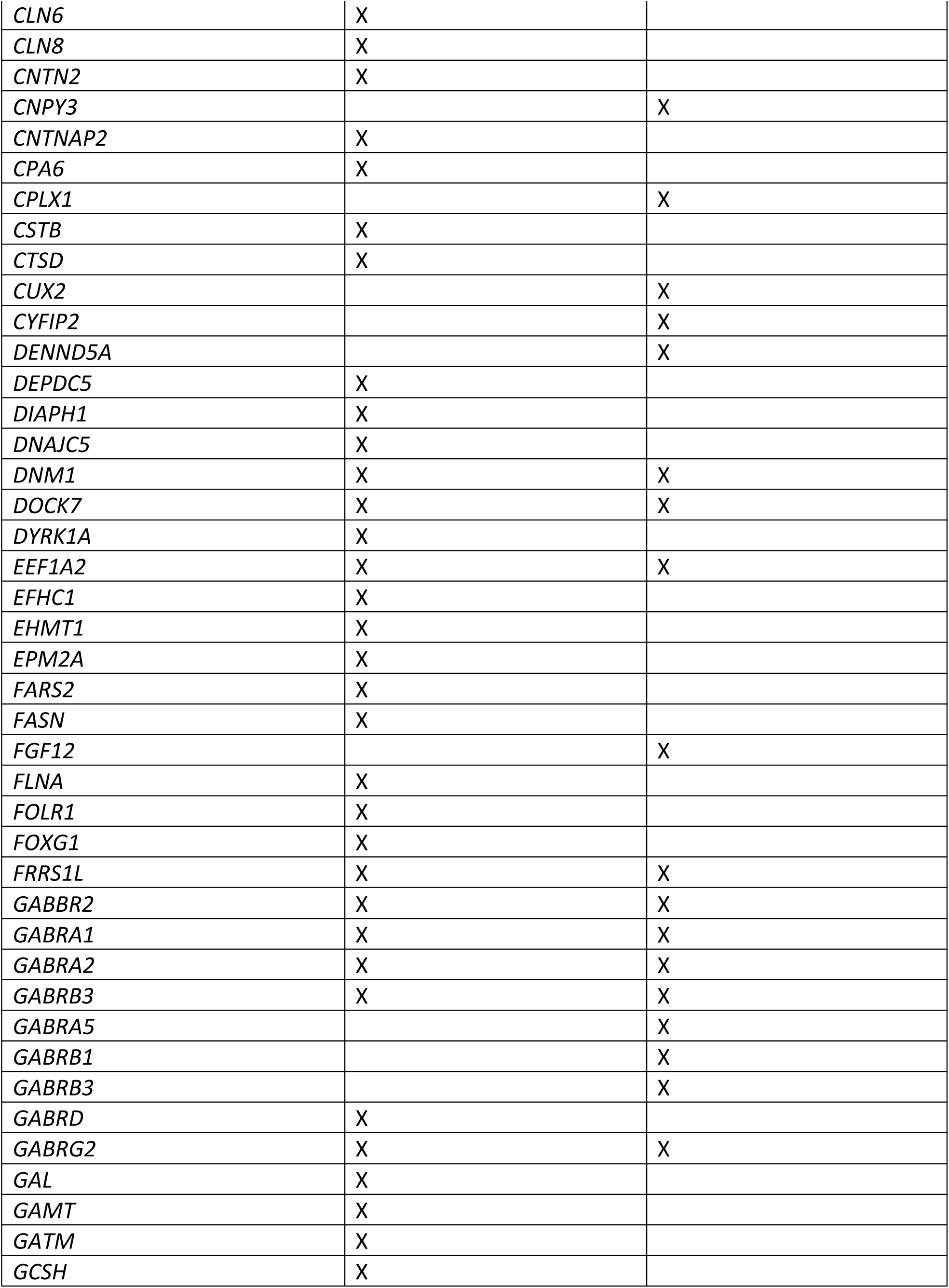
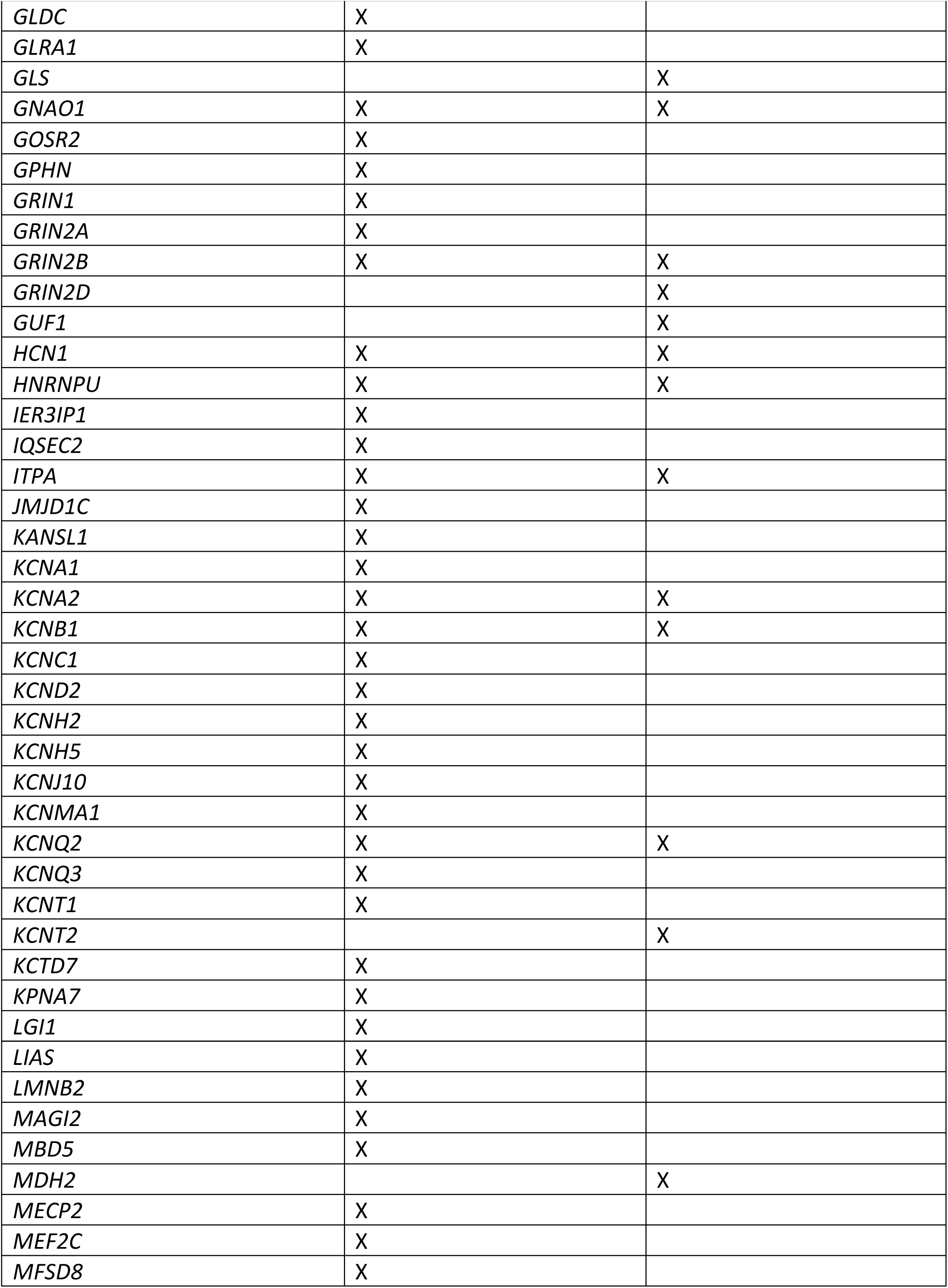
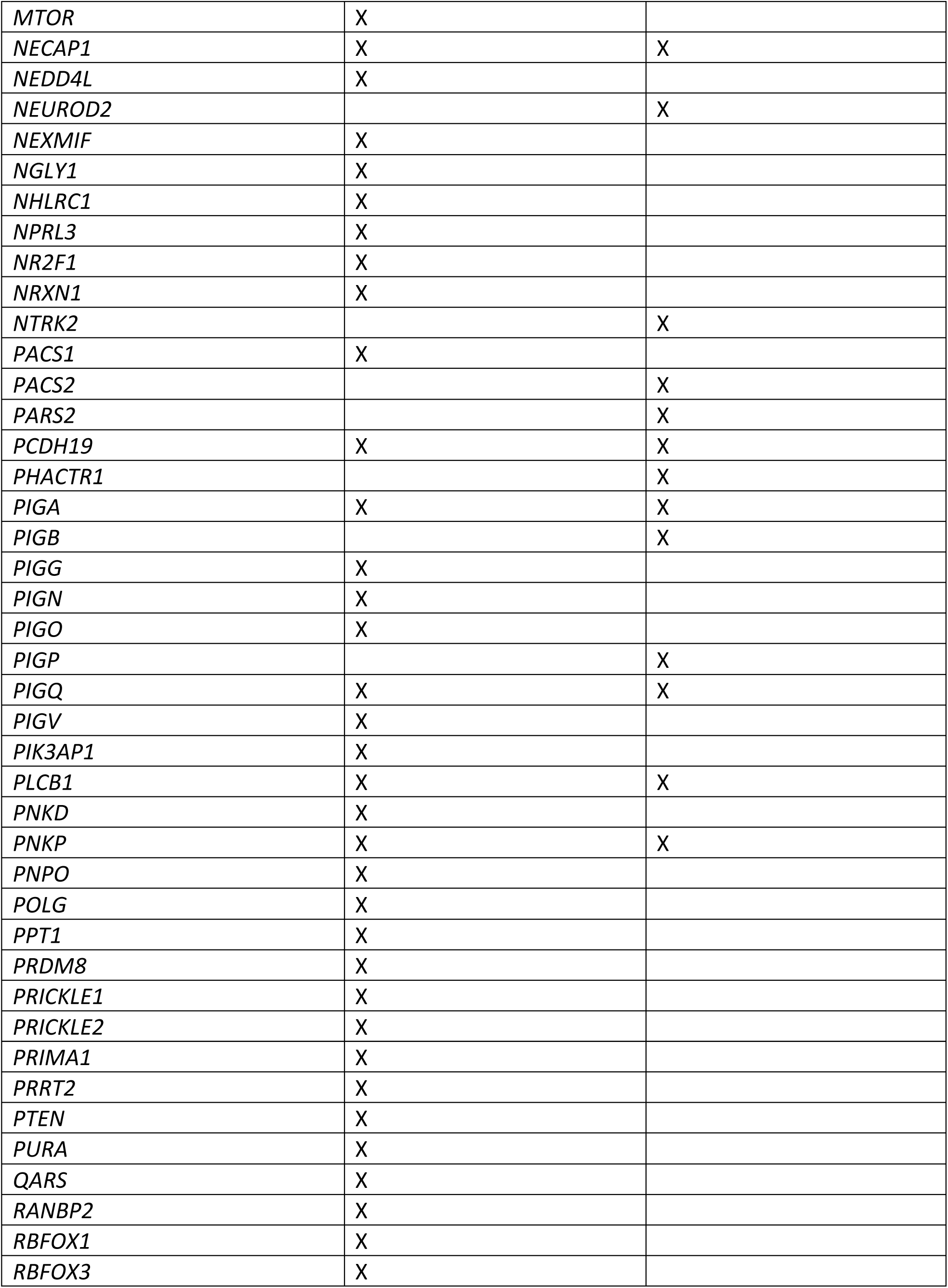
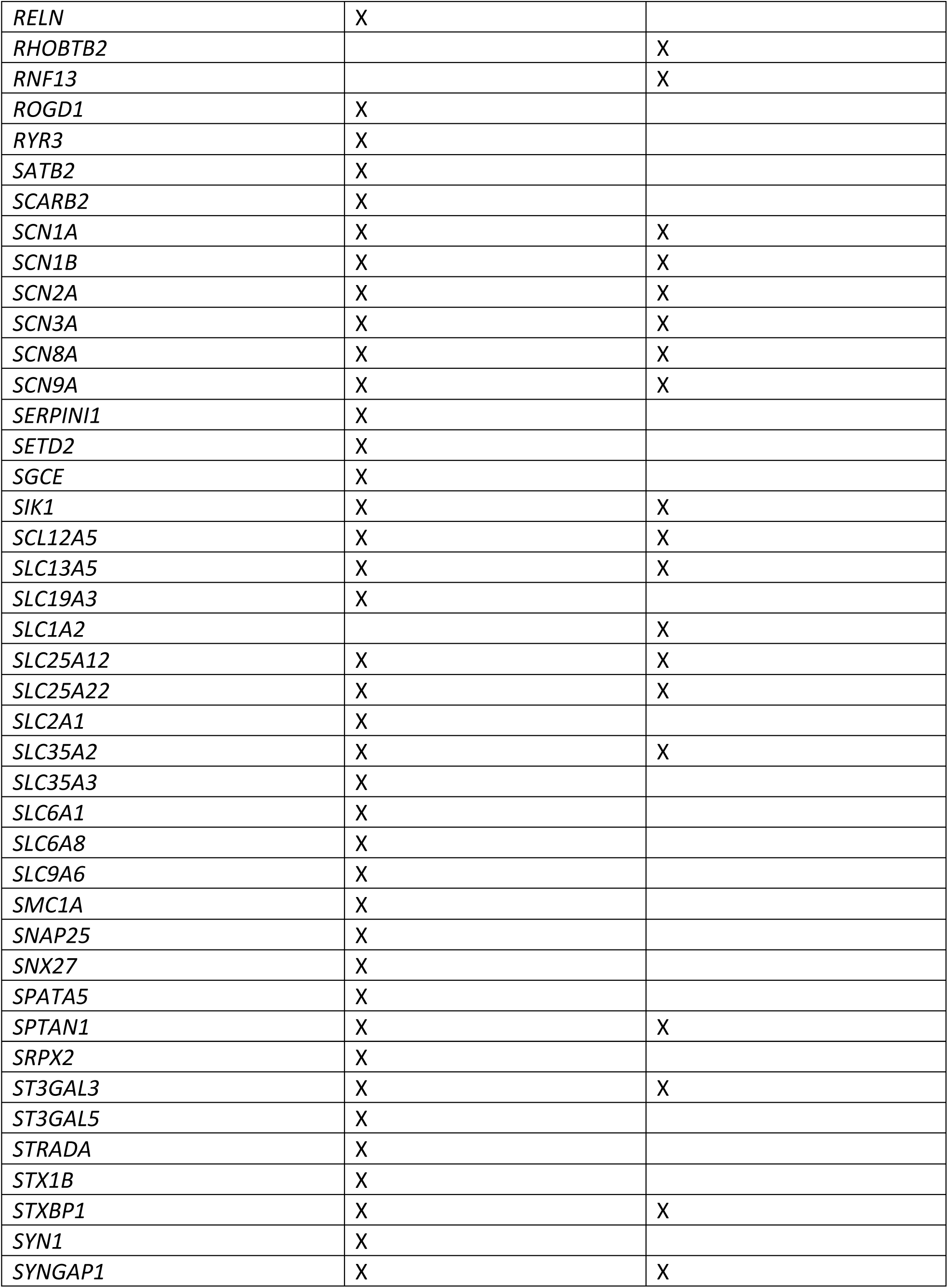
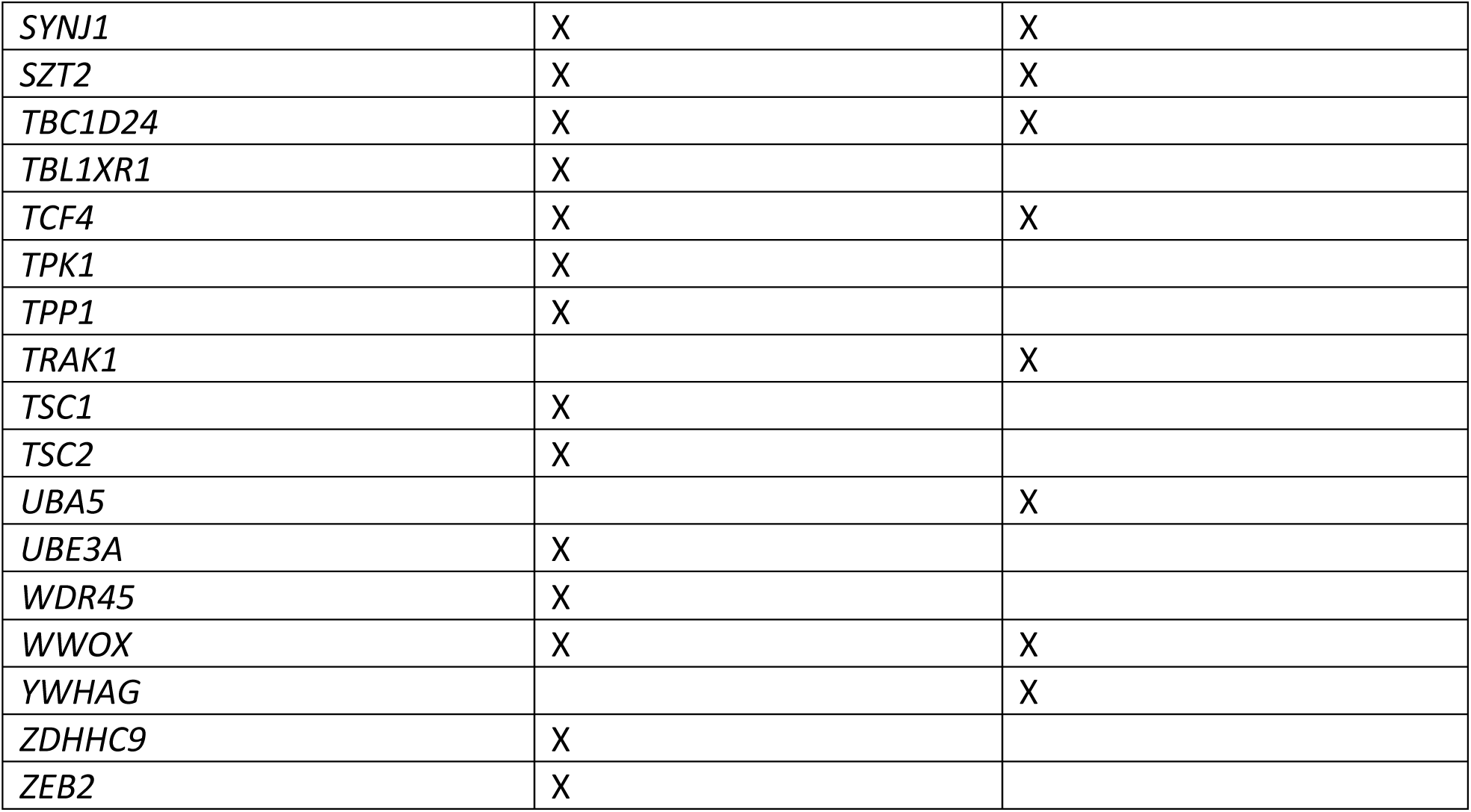
Epilepsy Gene panels

**Supplemental Table 3.** Cardiomyopathy and Arrhythmia Gene panels

| **Gene** | **CMAR1 n=118** | **CMAR2 n=143** |
| --- | --- | --- |
| *A2ML1* | X |  |
| *ABCC9* | X | X |
| *ACTC1* | X | X |
| *ACTN2* | X | X |
| *AGL* |  | X |
| *AKAP9* | X | X |
| *ALMS1* | X |  |
| *ANK2* | X | X |
| *ANKRD1* | X | X |
| *BAG3* | X | X |
| *BRAF* | X | X |
| *CACNA1C* | X | X |
| *CACNA2D1* | X | X |
| *CACNB2* | X | X |
| *CALM1* | X | X |
| *CALM2* | X | X |
| *CALM3* | X | X |
| *CALR3* |  | X |
| *CASQ2* | X | X |
| *CAV3* | X | X |
| *CBL* | X |  |
| *CHRM3* | X | X |
| *CRYAB* | X | X |
| *CSRP3* | X | X |
| *CTF1* | X | X |
| *CTNNA3* | X | X |
| *DEPDC5* |  | X |
| *DES* | X | X |
| *DMD* | X | X |
| *DOLK* | X | X |
| *DSC2* | X | X |
| *DSG2* | X | X |
| *DSP* | X | X |
| *DTNA* | X | X |
| *EMD* | X | X |
| *EYA4* | X | X |
| *FBN1* | X |  |
| *FHL1* | X | X |
| *FHL2* | X | X |
| *FKRP* | X | X |
| *FKTN* | *X* | *X* |
| *FLNC* | *X* | *X* |
| *GAA* |  | *X* |
| *GATA4* | *X* | *X* |
| *GATA6* | *X* |  |
| *GATAD1* | *X* | *X* |
| *GJA5* |  | *X* |
| *GLA* | *X* | *X* |
| *GPD1L* | *X* | *X* |
| *HCN4* | *X* | *X* |
| *HRAS* | *X* | *X* |
| *ILK* | *X* | *X* |
| *JPH2* | *X* | *X* |
| *JUP* | *X* | *X* |
| *KCNA5* |  | *X* |
| *KCND3* | *X* | *X* |
| *KCNE1* | *X* | *X* |
| *KCNE2* | *X* | *X* |
| *KCNE3* | *X* | *X* |
| *KCNE5* | *X* | *X* |
| *KCNH2* | *X* | *X* |
| *KCNJ2* | *X* | *X* |
| *KCNJ5* | *X* | *X* |
| *KCNJ8* | *X* | *X* |
| *KCNK3* |  | *X* |
| *KCNQ1* | *X* | *X* |
| *KCNQ2* |  | *X* |
| *KCNQ3* |  | *X* |
| *KCNT1* |  | *X* |
| *KRAS* | *X* | *X* |
| *LAMA4* | *X* | *X* |
| *LAMP2* | *X* | *X* |
| *LDB3* | *X* | *X* |
| *LMNA* | *X* | *X* |
| *LRRC10* |  | *X* |
| *MAP2K1* | *X* | *X* |
| *MAP2K2* | *X* | *X* |
| *MOG1* | *X* |  |
| *MTND1* |  | *X* |
| *MTND5* |  | *X* |
| *MTND6* |  | *X* |
| *MTTD* |  | *X* |
| *MTTG* |  | X |
| *MTTH* |  | X |
| *MTTI* |  | X |
| *MTTK* |  | X |
| *MTTL1* |  | X |
| *MTTL2* |  | X |
| *MTTM* |  | X |
| *MTTQ* |  | X |
| *MTTS1* |  | X |
| *MTTS2* |  | X |
| *MYBPC3* | X | X |
| *MYH6* | X | X |
| *MYH7* | X | X |
| *MYL2* | X | X |
| *MYL3* | X | X |
| *MYL4* | X | X |
| *MYLK2* | X | X |
| *MYOM1* | X | X |
| *MYOZ2* | X | X |
| *MYPN* | X | X |
| *NEBL* | X | X |
| *NEXN* | X | X |
| *NKX2-5* | X | X |
| *NPPA* |  | X |
| *NRAS* | X | X |
| *PCDH19* |  | X |
| *PDLIM3* | X | X |
| *PKP2* | X | X |
| *PLEKHM2* |  | X |
| *PLN* | X | X |
| *PRDM16* | X | X |
| *PRKAG2* | X | X |
| *PRRT2* |  | X |
| *PTPN11* | X | X |
| *RAF1* | X | X |
| *RANGRF* |  | X |
| *RASA1* | X |  |
| *RBM20* | X | X |
| *RIT1* | X |  |
| *RYR2* | X | X |
| *SCN10A* | X | X |
| *SCN1A* |  | X |
| *SCN1B* | *X* | *X* |
| *SCN2B* | *X* |  |
| *SCN3B* | *X* | *X* |
| *SCN4B* | *X* | *X* |
| *SCN5A* | *X* | *X* |
| *SCN8A* |  | *X* |
| *SCN9A* |  | *X* |
| *SDHA* | *X* |  |
| *SGCD* | *X* | *X* |
| *SHOC2* | *X* |  |
| *SLC22A5* |  | *X* |
| *SLC2A1* |  | *X* |
| *SLMAP* |  | *X* |
| *SNTA1* | *X* | *X* |
| *SOS1* | *X* | *X* |
| *SPRED1* | *X* |  |
| *TAZ* | *X* | *X* |
| *TCAP* | *X* | *X* |
| *TGFB3* | *X* | *X* |
| *TMEM43* | *X* | *X* |
| *TMPO* | *X* | *X* |
| *TNNC1* | *X* | *X* |
| *TNNI3* | *X* | *X* |
| *TNNT2* | *X* | *X* |
| *TPM1* | *X* | *X* |
| *TRDN* | *X* | *X* |
| *TRPM4* |  | *X* |
| *TTN* | *X* | *X* |
| *TTR* | *X* | *X* |
| *TXNRD2* | *X* | *X* |
| *VCL* | *X* | *X* |

**Supplemental Table 4.** Pathogenic and Likely Pathogenic Variants Identified in Epilepsy and Cardiac Genes

| ID* | Age<br>at<br>Death<br>(yrs) | Cause of<br>Death | ClinVar | GENE | gDNA | AA<br>Change | gnomAD |
| --- | --- | --- | --- | --- | --- | --- | --- |
| **<u>Epilepsy Panel Gene Variants</u>** |  |  |  |  |  |  |  |
| 1010 | <1 | Cardiac,<br>congenital | LP | QARS* | 3:49137655<br>G>A | p.Arg367Cys | 3.19E-05 |
| 1172 | <1 | Infant<br>Suffocation | LP | CLN8* | 8:1719428<br>C>T | p.Arg70Cys | 0 |
| 1193 | 10-20 | Venous<br>Malformation | P | PPT1* | 1:40558081<br>T>G | p.Thr25Pro | 0 |
|  | 10-20 | Venous<br>Malformation | P/LP | CSTB* | 21:45194641<br>C>G | n.104G>C | 0.0003 |
| **<u>CMAR1 Panel Gene Variants</u>** |  |  |  |  |  |  |  |
| 1001 | <1 | Unexplained | P | FKRP* | 19:47259533<br>C>A | p.Leu276Ile | 0.0014 |
| 1012 | <1 | Unexplained | P | TTR | 18:29178618<br>G>A | p.Val142Ile | 0.0048 |
| 1028 | <1 | Unexplained | P | TTR | 18:29178618<br>G>A | p.Val142Ile | 0.0048 |
| 1058 | >1-<br><10 | Unexplained | LP | CALM3 | 19:47112212<br>A>G | p.Asp96Gly | 0 |
| 1117 | <1 | Infant<br>Suffocation | P | TTR | 18:29178618<br>G>A | p.Val142Ile | 0.0048 |
| 1127 | <1 | Unexplained | P | TTR | 18:29178618<br>G>A | p.Val142Ile | 0.0048 |
| 1132 | NA | NA | P | DSP | 6:7571745<br>C>T | p.Gln611* | 0 |
| **<u>Epilepsy and CMAR1 Panel Gene Variants</u>** |  |  |  |  |  |  |  |
| 1214 | NA | NA | P | KCNH2 | 7:150654393<br>C>CAG | p.Thr371_Glu372fs | 0 |

676\*All IDs are created for WGS purposes and not to reidentify subjects. Variants in this table were 677annotated with population frequency, protein effect, literature review, and *in silico* modeling. 678Pathogenicity was determined by methods similar to ACMG standards (see Methods). NA= Not 679reported in case review. P=Pathogenic; LP=Likely Pathogenic. \*Gene is generally associated with 680autosomal recessive inheritance and decedent is heterozygous. AA= amino acid

**Supplemental Table 5.**
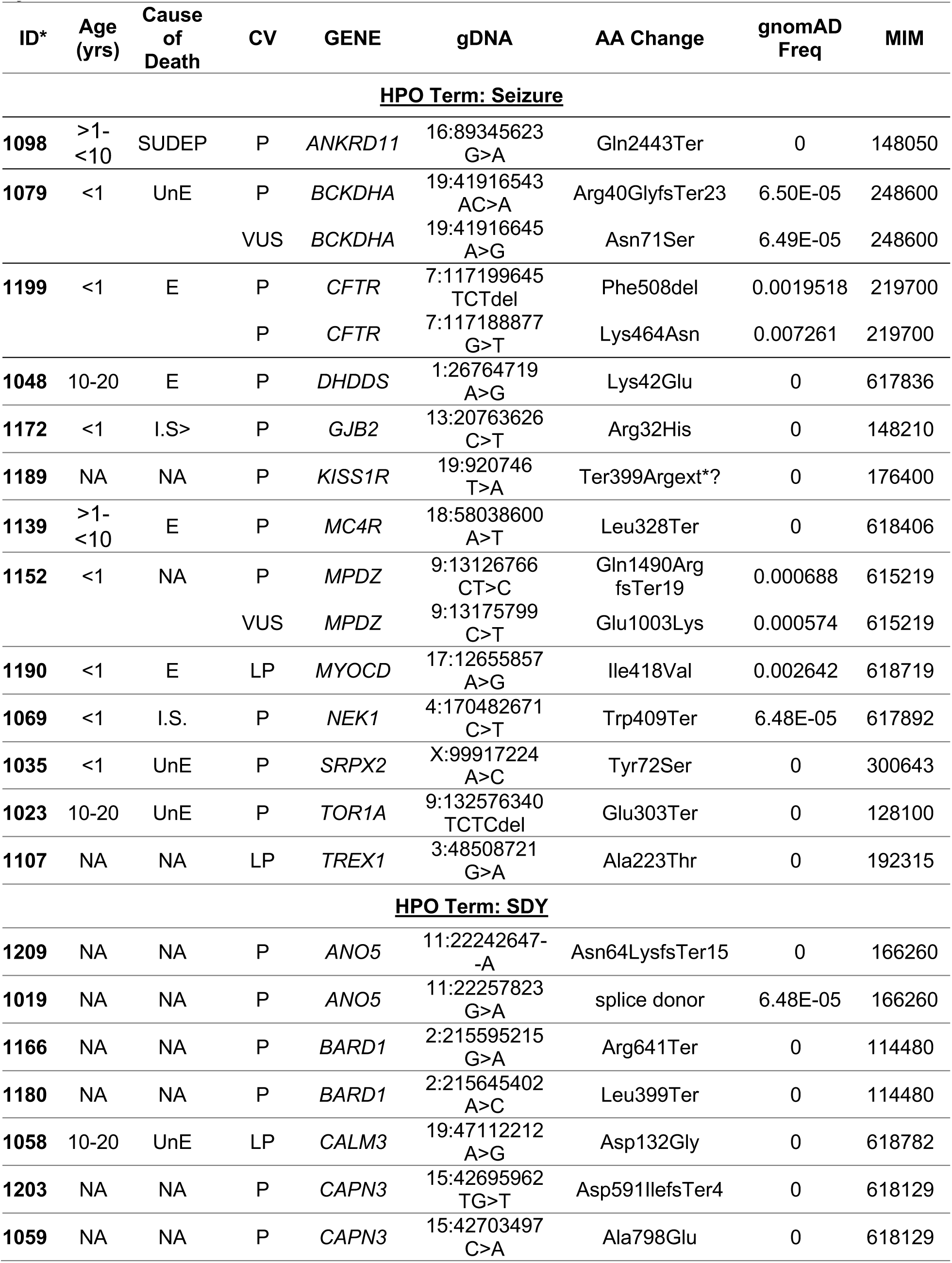
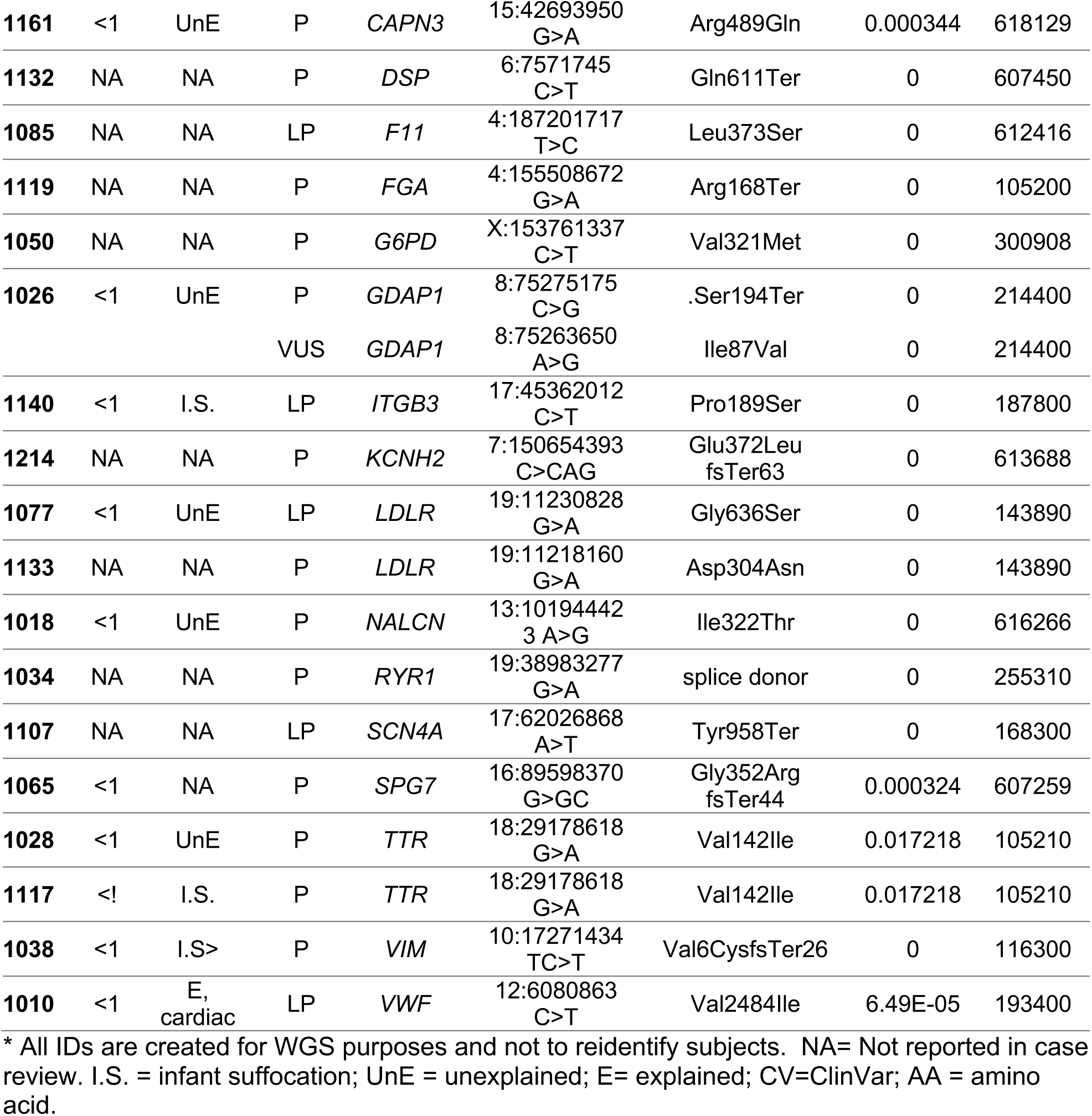
Pathogenic and Likely pathogenic, Mendelian variants as ranked by GEMS

**Supplemental Table 6.** Enrichment of variants in epilepsy or cardiac genes in the SDY cohort compared to a ancestry and sex matched 1000 genomes cohort.

| GENE LIST | PHENOTYPE TERMS |  |  |  |
| --- | --- | --- | --- | --- |
|  | Seizure |  |  |  |
|  | SDY | 1000 Genomes Project | Genes | Adjusted p-Value* |
| **Epilepsy** | 62 | 36 | 191 | **0.027** |
| **EIEE-OMIM** | 38 | 16 | 82 | **0.019** |
|  | Cardiac |  |  |  |
| **CMAR1** | 55 | 27 | 118 | **<0.001** |
| **CMAR2** | 58 | 31 | 143 | **<0.001** |

\*P values adjusted for multiple comparisons using the FDR adjustment.

**Supplemental Table 7.** Linear regression of age at death against number of rare epilepsy
variants*.

| **Model** | **Coefficient** | **P value** |
| --- | --- | --- |
| Adjusted (Ancestry PCs 1-6) | -0.647 | 0.0053 |
| Unadjusted | -0.458 | 0.0387 |
| Adjusted log | -0.159 | 0.0494 |
| ***Extreme value sensitivity analysis*** |  |  |
| Removal of cases with more than 9 variants (Adjusted PCs) | -0.636 | 0.0113 |
| Removal of cases with more than 9 variants and less than 1 (Adjusted PCs) | -0.638 | 0.0137 |
| Removal of cases with 0 rare epilepsy(<0.001) variants (Adjusted PCs) | -0.650 | 0.0063 |

\*Rare epilepsy variants = nonsynonymous variants identified in the Epilepsy gene panel with an allele frequency <0.001 in gnomAD.

